# Sex-specific effects of Birth Weight on Longitudinal Behavioural Outcomes; a Mendelian Randomisation Approach using Polygenic Scores

**DOI:** 10.1101/2024.01.22.24301633

**Authors:** Lars Meinertz Byg, Carol Wang, John Attia, Andrew Whitehouse, Craig Pennell

**Author notes:** **Corresponding author:** Lars Meinertz Byg, MD, Mailing address: Lot 1 Kookaburra Cct, New Lambton Heights NSW 2305.

## Abstract

**Intro:** It is unclear if sex differences in behaviour arising from birth weight (BW) are genuine because of the cross-sectional nature and potential confounding in previous studies. We aimed to test if sex differences associated with birth weight phenotype were reproducible using a Mendelian randomisation approach, i.e. polygenic score for birthweight across childhood and adolescence.

**Method:** Utilising data from the Raine study we had 1484 genotyped participants with a total of 6446 child behaviour checklist assessments across childhood and adolescence. We used BW polygenic scores in linear mixed-effects models to predict parentally-assessed attention, aggression and social problems scales; we also derived estimates and significance for a sex-by-genotype interaction. We used a Bonferroni corrected significance threshold and tested robustness of the results with teacher assessments of behaviour as well as a second polygenic score.

**Results:** We found a sex-by-genotype interaction with lower BW polygenic scores (BW-PGS) associated with increased aggression in males compared to females. These findings were consistent across various analyses, including teacher assessments. Surprisingly, a lower BW-PGS showed protective effects in females, while lower BW phenotype had detrimental effects in males with evidence of a genotype-phenotype mismatch increasing aggression problems in males only.

**Conclusion:** This study underscores the genuine nature of behavioural sex differences arising from low BW and highlights the sex-dependent and diverging effects of environmental and genetic BW determinants.

## Introduction

Mental illness is believed to be caused by gene-environment interactions and often begins in early childhood [1]. Birth weight (BW) is associated with psychopathology and multiple diagnostic categories [2–4]. Similarly, childhood behaviour has been associated with BW in multiple settings [5–8] and Mendelian randomisation has provided evidence that low BW increases the risk of mental illness [9]. Translational studies on developmental programming support these conclusions but also find sex differences in programmable behaviour outcomes [10, 11].

Sex differences in psychiatric epidemiology are well-established [12] and sex may also modify the behavioural effects of BW [13–15], i.e. despite females having generally lower BW, sex may independently affect behaviour. The largest study to date found that low BW increased attention and aggression problems only in males with a non-significant signal for social problems at school age [13]; however, another study reported that females with lower BW were at increased risk for attention problems at preschool age [14]. Intriguingly, both low BW and biological sex are implicated as determinants of behavioural phenotype trajectories (longitudinal outcomes) across childhood and adolescence [16, 17]. This raises the possibility that biological sex does not change the behavioural phenotypes induced by low BW per se but simply the age at which these effects are manifested. Additionally, measured BW is a variable with a substantial risk of residual confounding [18]. Taken together with the inconsistencies in previous studies, causal inferences on lasting sex differences in behavioural phenotypes from lower BW, and potential mechanisms for this, are premature.

Both genes and environment during gestation affect BW, and sex differences from low BW may stem from sex-by-environment or sex-by-gene interactions [19]. To our knowledge, no twin study has tested the sex differences in behavioural outcomes from low BW; however, a sibling control design in Sweden found similar effect sizes of low BW on diagnostic rates of psychiatric illnesses for concordant male and female sibling pairs [20]. The contrast between this sibling and the non-related population studies cited above may be explained by confounding or the use of lifetime diagnoses versus continuous behavioural assessments. Alternatively, behavioural outcomes from low BW may only show sex differences in more genetically diverse settings. Taken together, the evidence suggests that repeated measures across childhood and adolescence are needed to distinguish if genetic determinants of BW drive lasting behavioural sex differences seen in low BW.

Our study sought to investigate if a Mendelian randomisation approach using polygenic scores for birth weight had different effects on behaviour in males and females across childhood and adolescence focusing on attention-, aggression- and social problems; this approach should be free of traditional confounders and allow more robust inferences about causation.

## Methods

### Study population

We analysed data from the Raine Study (https://rainestudy.org.au/)[21]. The Raine Study is a longitudinal study following mother-baby dyads recruited at or around 18 weeks gestation (n=2979) through the public antenatal clinic at King Edward Memorial Hospital and nearby private clinics in Perth, Western Australia, from May 1989 to November 1991. Generation 2 participants were followed up throughout childhood with modest attrition of mothers with lower age, education, income and non-European ancestry[22, 23]. The Human Research Ethics Committees at the University of Western Australia, King Edward Memorial Hospital, and Princess Margaret Hospital in Perth, Australia, granted ethics approvals. Participants were eligible for analysis if they had data available on biological sex, at least one follow up behavioural assessment, and genetic data.

### Childhood behavioural assessments

Using parent reports of childhood behaviour checklist (CBCL) for Ages 4–18 (CBCL/4–18), we derived scores of attention- (scale from 0 – 22), aggression- (scale from 0-40) and social problems (scale from 0-16) at ages five, eight, ten, 14 and 17. These three behavioural measures were chosen based on the report by Dooley et al who found sex differences in a cohort with similar ancestry as ours (American versus Australian)[13]. The CBCL/4–18 is a commonly used dimensional measure of child behaviour during the previous six based on parent ratings of 118 items on a 3-point Likert scale (0= “Not true” 1 = “somewhat or sometimes true”, 2 = “Very often or often true”)[24]. For aggression problems, we also had available measures from the ASEBA preschool form from two-three years of age, included in sensitivity analysis. The 1991 edition of the preschool questionnaire did not have attention problem and social problem scale and the aggression score had more items (scale from 0-66) than the CBCL/4-18 [25]. Assessments were excluded from analysis if they were missing more than 8 items on the entire CBCL [26]. The attention problem subscale measures both problems of attention, impulsivity and hyperactivity and consists of 11 items. The social problems scale measures problems of peer interaction and consists of eight items in total. The aggressive behaviour scale measures both direct and relational aggression and consists of 20 items. The CBCL is a highly validated, clinically used, psychometric tool available in many countries [24, 27, 28]. The teacher report form (TRF) is another form by the same provider, used in the clinical setting to support findings from the CBCL[29]. The TRF also has an attention- (20 items, score 0-40), aggression- (25 items, score 0-50) and social problems scale (13 items, score 0-26).

### Development of the birth weight polygenic score

Information on Raine study genotyping and quality control is provided in the supplementary materials. Genome-wide association analyses have enabled us to identify individual genetic variants associated with a wide range of traits; however, these effect sizes are usually small with low predictive power [30]. Several studies demonstrated increased predictive power with the use of polygenic score (PGS), a metric computed by summing the risk alleles associated with the phenotype of interest in each individual, when compared to a small number of genome-wide significant SNPs [31, 32]. In addition to identifying and understanding potential aetiology of disease, PGS have also been used to test for genome-wide Gene-Gene and Gene-Environment interactions [31, 33]. To simplify interpretation of the BW-instrument, we identified 146 SNPs associated with offspring BW (i.e. effects on BW through the fetus rather than the mother)[34]. These SNPs data were extracted from the Raine Study Gen2 participants, re-coded to correspond with increasing BW, weighted using the beta-coefficients reported in the meta-analysis, summed and re-scaled before the BW-PGS was calculated for each study participant. The final BW-PGS was then normalised to facilitate comparison with measured BW.

### Early life determinants and potential confounders

Measured BW and fetal sex were retrieved from hospital records. For one participant with missing data we extracted sex from a later questionnaire. The recorded BW was normalised for analysis to ease comparison with the BW-PGS. Preterm birth was defined as gestational age (GA) < 37 weeks, and GA was determined by either date of last menstrual period (LMP) or fetal biometry at the 18-week gestation ultrasound (USS) examination.

### Statistics

All statistical analyses and graph productions were performed in R and its associated libraries “ggplot2”, “lme4”, “lmeresampler”, and “lmertest” [35]. Wilcoxon rank sum test was used to compare sample distributions in the continuous baseline variables and CBCL outcomes between males and females.

In order to use the repeated behavioural measures most efficiently, we used linear mixed-effects modelling to test the associations between BW-PGS and behavioural outcomes; a random effect was used for participant ID and fixed effect for the BW-PGS and other co-variables. Recent work has demonstrated that the treatment of ordinal data as continuous does not impact inference in most situations [36], and linear mixed modelling is robust to missing data and assumption violations [37]. We treated the CBCL scores as continuous variables due to the dimensional nature of psychopathology[38]. The clinical validation of the CBCL is always done with adjustment for age and sex[27], and so the grouping variable for age at assessment was included in the regression giving us two models:

Model 1 containing fixed effects of the BW-PGS and age at assessment, and
Model 2 with the additional fixed effect for sex and a sex x PGS interaction (see Figure 1 for directed acyclic graph).

**Figure 1.**
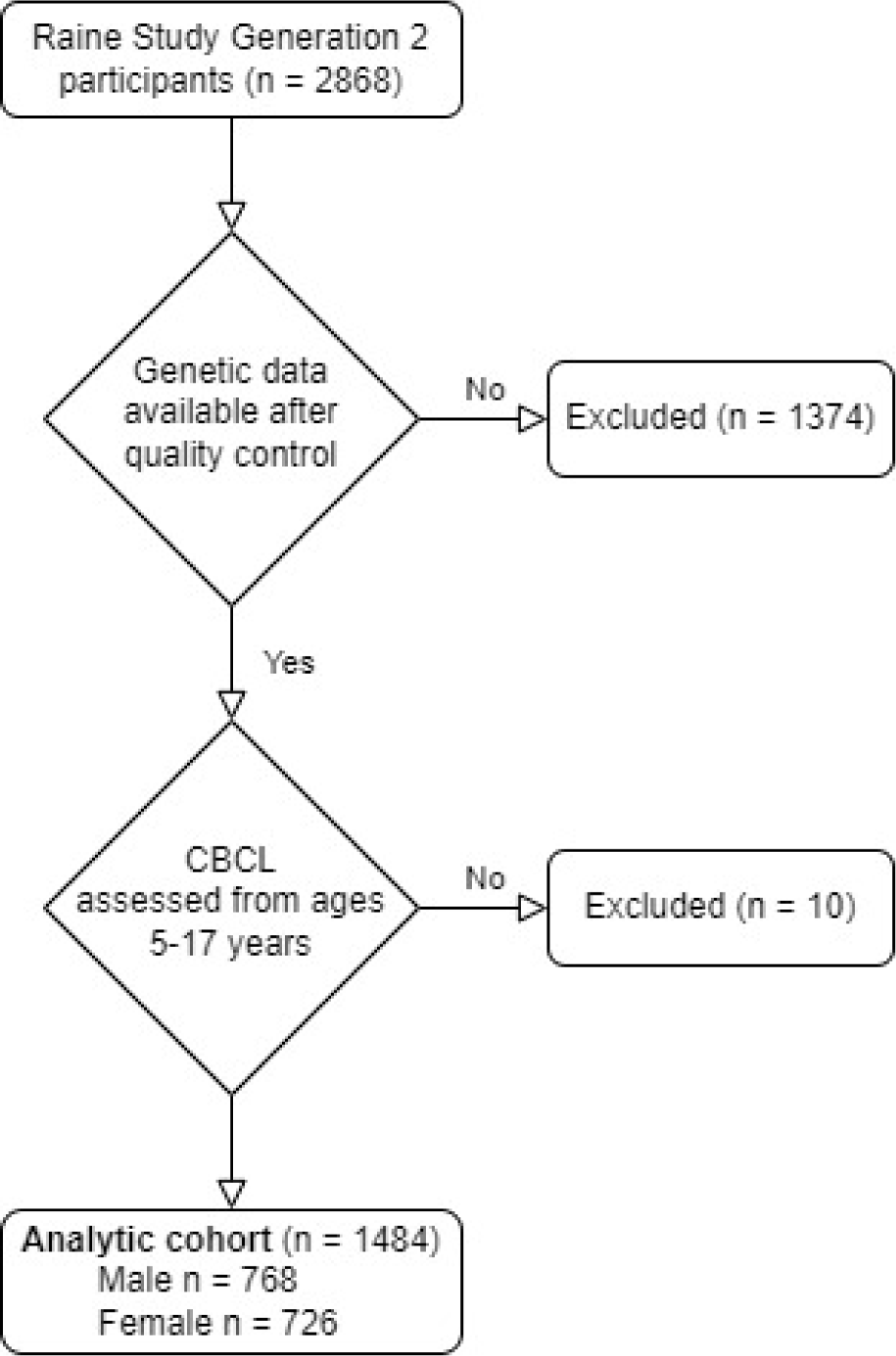
Flow diagram of cohort selection. **CBCL:** Child behaviour checklist

**Figure 2.**
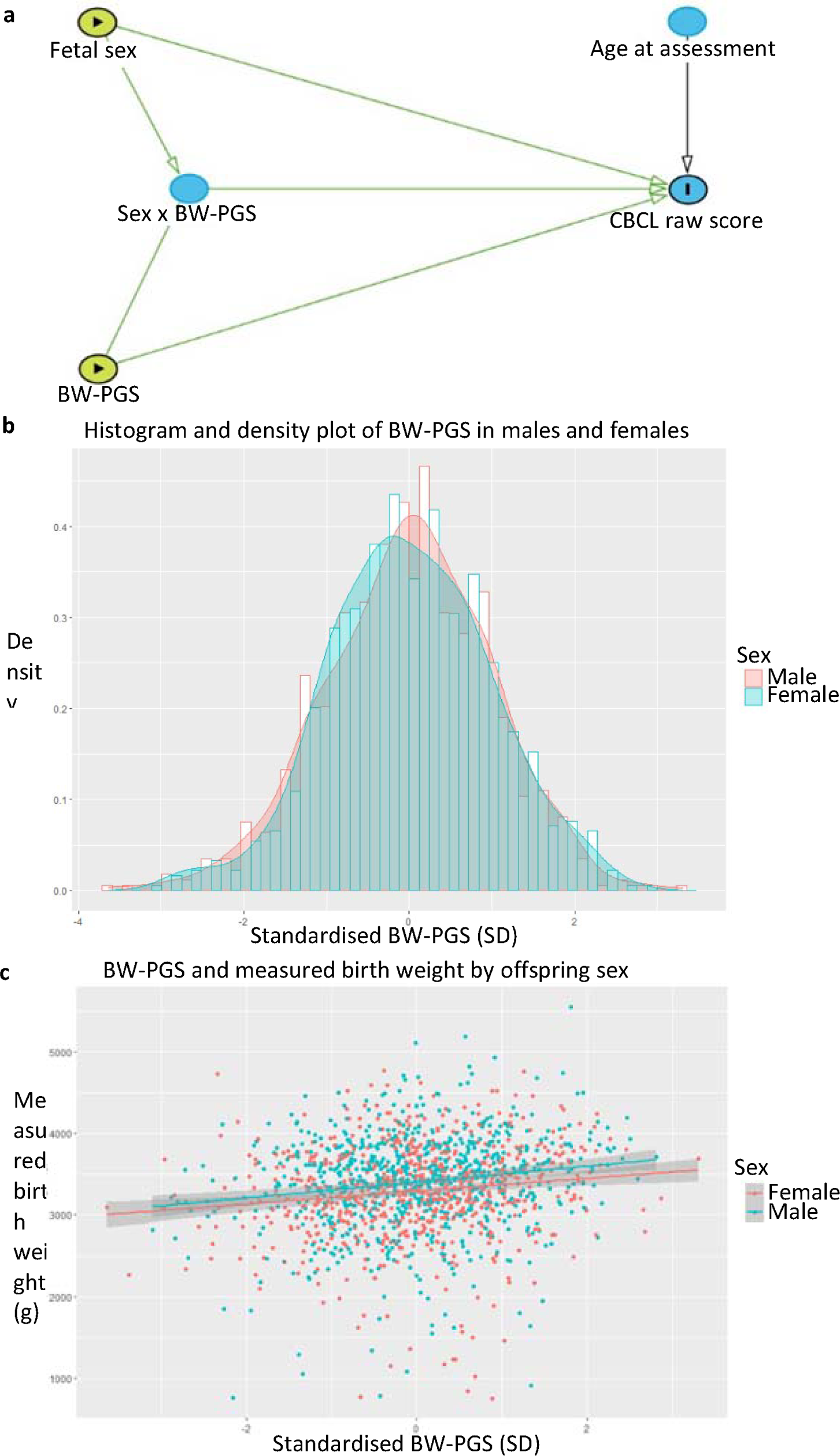
Directed acyclic graph and birth weight polygenic score. Conceptual framework for the primary analysis demonstrated in a Directed Acyclic Graph (DAG) (**a**). Density plot and histogram of the distribution of our birth weight polygenic scores in males and females (**b**) and the effect of the polygenic score on measured birthweight in males and females (**c**)

If *y_IA_* is the CBCL score for a participant ID at a specific age of assessment, *B*_0_ is the grand mean intercept, *u*_*I*0_ is our ID-random effect on the intercept, *B*_1_ is the coefficient estimate of *PCS_IA_* in females, ***B***_2_ is the interaction of male sex on *B*_1_, *B*_3_ is the fixed effect of age, and *B*_4_ is the fixed effect of sex, and finally *ε_IA_* is the error term, our regression equation for model 2 is:

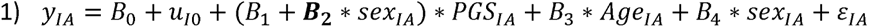

The effect estimate of the sex-interaction term in bold **B**_2_ was the variable of interest for our primary research question. Due to our interest in three different behavioural outcomes, we applied a Bonferroni correction yielding a significance level of 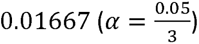 and the corresponding 98.3 % confidence interval was used for primary inference. To investigate the similarity in outcome-exposure relationships between our cohort and previous studies, we supplemented our analyses by running our model two using the measured BW in the genotyped cohort as the exposure with adjustment for age at assessment and biological sex.

Models diagnostics were evaluated by histograms and qq-plots of residuals and random effects. After finding assumption violations in the quality control, a non-parametric bootstrap at the participant-ID level with 3000 simulations was used to derive SE and 98.3 % confidence intervals as suggested by Thai et al. [39].

As sensitivity analyses, we reanalysed our model with term-born offspring only, and with the inclusion of an aggression assessment at age two. Parents have clear associations with both the BW-PGS and CBCL, and we therefore wanted to ensure the directionality of our findings with an independent analysis of teacher assessments at age ten. This was done with linear regression and case-bootstrapping of confidence intervals with 3000 simulations.

After finding large effect sizes for the BW-PGS, we reanalysed our models using an earlier BW-PGS (BW-PGS2) [40] and added primary components to examine confounding from population stratification. After finding that non-genetic (i.e. environmental) factors account for the majority of variance in birth weight[19], we sought to test if there was an interaction between BW phenotype (reflecting environmental determinants of BW) and BW genotype (reflecting genetic determinants of BW) in determining behaviour. BW-PGS is causal for measured BW and to avoid collinearity and bias in the interaction term we generated a new variable “environmental BW” (BW-ENV and BW-ENV2 for BW-PGS and BW-PGS2, respectively) by using the variance from a linear regression of measured BW on BW-PGS and BW-PGS2. We then tested for a genotype-phenotype interaction (“BW-PGS x BW-ENV” and “BW-PGS2 x BW-ENV2”) using a linear mixed model with inclusion of fixed effects for BW-PGS and BW-ENV for males and females separately and together.

## Results

The analytic cohort for the association between BW-PGS and behaviour consisted of 1484 participants with outcomes for ages five to 17, corresponding to 52 % of the original Gen2 Raine Study participants (Figure 1), with a total of 6446 CBCL assessments over the follow-up period (supplementary Table 1). The maternal baseline characteristics were similar between male and female fetuses (supplementary Table 2). The genotyped cohort displayed selective attrition on a range of maternal baseline characteristics compared to the original Raine Study cohort, including lower BW (Supplementary Table 3).

In our analytic cohort, males had significantly higher BW than females (p < 0.0001) and males generally had higher CBCL scores on the subscales of interest. In the genotyped cohort, males and females had a similar BW-PGS (p = 0.8), and the BW-PGS was normally distributed (Supplementary Table 1 and Figure 1). Without stratification for sex, a 1-SD increase in PGS resulted in an 88.9 g increase in BW (p < 0.0001), explaining 2.25 % of the BW variance. This relationship had no significant sex interaction (p = 0.54) (Figure 1). Analysis of the behavioural outcomes using measured BW (n = 1483) suggested a sex-interaction where males had more behavioural problems at lower BW as compared to females, but this interaction did not reach statistical significance (all interaction p-values > 0.0167) (supplementary Table 4)

We then moved on to the primary research question regarding sex differences in the longitudinal behavioural effects of the BW-PGS (Table 1). We found no main effect of the BW-PGS on aggression problems, attention problems or social problems (p-value for all > 0.0167). Second, we examined the regression output for the effects of the BW-PGS on behavioural outcomes in males and females separately. For aggression problems in females, a 1SD increase in BW-PGS resulted in a 0.41 (**98.3%CI: [0.069, 0.744], P-value: 0.0038**) point increase in CBCL aggression problems score, whereas in males a 1 SD increase in BW-PGS did not significantly affect aggression problems (***β***: −0.171, 98.3% CI: [−0.534, 0.188], *P-value: 0.259) (Figure 3). The difference between female and male effects is significant given the formal interaction term was significant (**p=0.0045**). For attention problems in females, a 1 SD increase in the BW-PGS resulted in a 0.179 (**98.3% CI: [0.019, 0.331], P-value: 0.0063**) point increase in CBCL attention problems score, whereas in males a 1 SD decrease in BW did not significantly reduce the effect of BW-PGS on attention problems (***β***: 197 cl:13590.00907, 98.3% CI: [−0.184, 0.158], P-value: 0.899). This difference between females and males was not significant at our Bonferroni corrected threshold (p = 0.0498). For social problems, we found no significant effect of BW-PGS in females (***β***: 0.077, 98.3% CI: [−0.0488, 0.198], P-value: 0.138) or males (***β***: 0.033, 98.3% CI: [−0.068, 0.130], P-value: 0.421.) and the interaction term was not significant (p=0.52)

**Figure 3.**
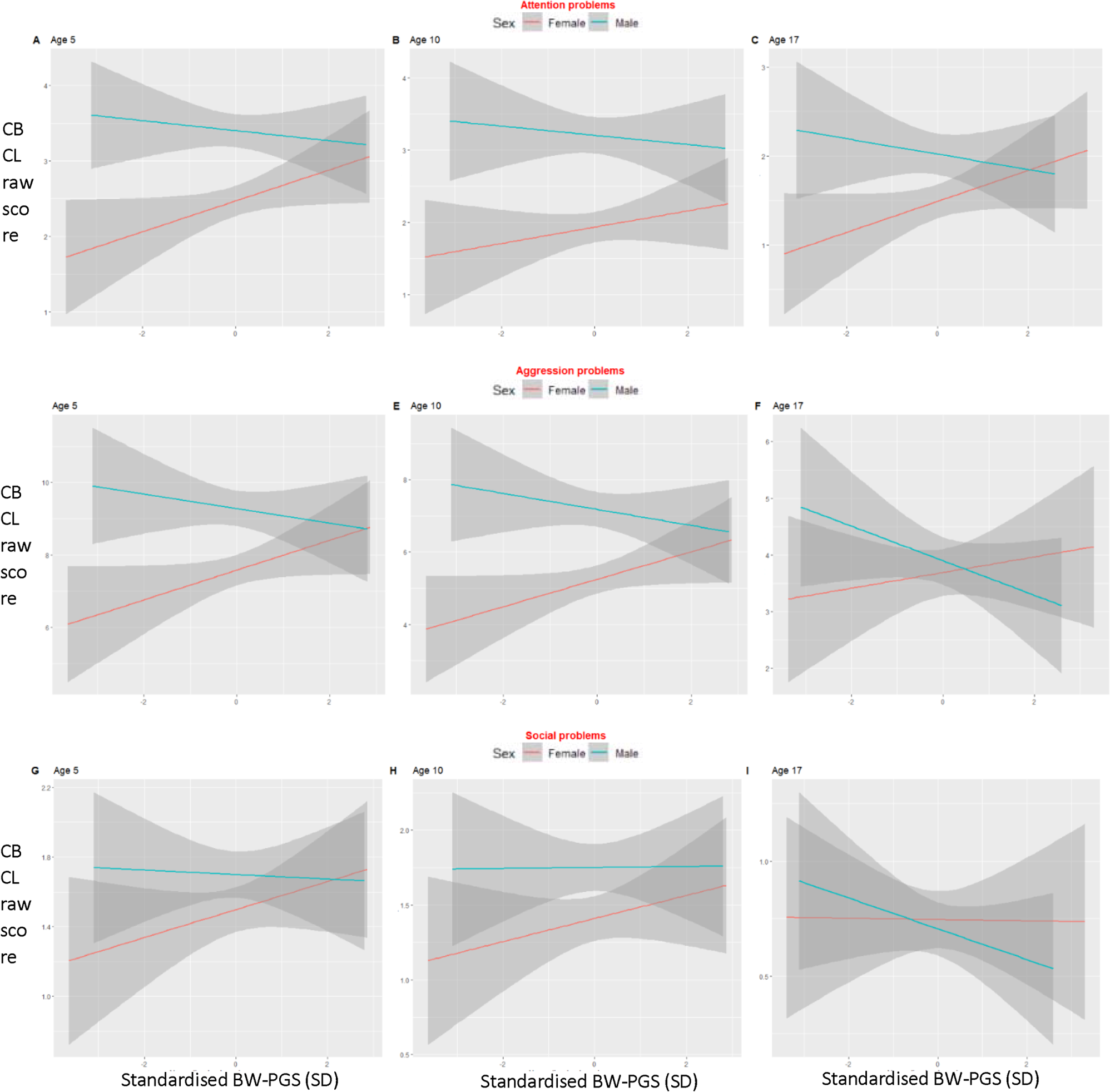
Graphical illustration of the sex interaction on behavioural effects of birth weight genetics. The linear relationship between the normalised PGS and attention- (**A-C**), aggression- (**D-F**) and social problems(**G-I**) at ages five, ten and 17 in the Raine Study in males and females. Note that the Y-axis changes depending on the CBCL syndrome and the age at assessment. **BW-PGS:** Birth weight polygenic score, **CBCL:** Child behaviour checklist.

**Table 1.** Primary analysis output of interaction between sex and birth weight genotype. Primary analysis showing the regression output for our linear mixed effects model. * p-value was calculated based on the z-statistic from the bootstrapped standard error, and the CI represents the 98.3% confidence interval.

|  | Aggression problems |  | Attention problems |  | Social problems |  |
| --- | --- | --- | --- | --- | --- | --- |
|  | Model 1 | Model 2 | Model 1 | Model 2 | Model 1 | Model 2 |
| Main effect (BW-PGS) | $\beta$: 0.117<br>*CI: [-0.139, 0.363]<br>SE: 0.105<br>*P-value: 0.266 | NA | $\beta$: 0.085<br>*CI: [-0.034, 0.200]<br>SE: 0.05<br>*P-value: 0.083 | NA | $\beta$: 0.055<br>*CI: [-0.026, 0.130]<br>SE: 0.033<br>*P-value: 0.091 | NA |
| Female effect | NA | $\beta$: 0.410<br>*CI: [0.069, 0.744]<br>SE: 0.141<br>*P-value: 0.0038 | NA | $\beta$: 0.179<br>*CI: [0.019, 0.331]<br>SE: 0.065<br>*P-value: 0.0063 | NA | $\beta$: 0.077<br>*CI: [-0.049, 0.198]<br>SE: 0.052<br>*P-value: 0.138 |
| Male effect | NA | $\beta$: -0.171<br>*CI: [-0.534, 0.188]<br>SE: 0.151<br>*P-value: 0.259 | NA | $\beta$: -0.00907<br>*CI: [-0.184, 0.158]<br>SE: 0.0717<br>*P-value: 0.899 | NA | $\beta$: 0.033<br>*CI: [-0.068, 0.130]<br>SE: 0.0416<br>*P-value: 0.421 |
| Interaction on female effect from male sex | NA | $\beta$: -0.581<br>*CI: [-1.06, -0.089]<br>SE: 0.204<br>*P-value: 0.0045 | NA | $\beta$: -0.188<br>*CI: [-0.416, 0.041]<br>SE: 0.096<br>*P-value: 0.0498 | NA | $\beta$: -0.043<br>*CI: [-0.203, 0.117]<br>SE: 0.067<br>*P-value: 0.52 |

As a prespecified sensitivity analysis, we included the preschool aggression problems assessment, yielding an additional 5 participants for analysis (n = 1489). Inclusion of this assessment was consistent with the primary analysis (Supplementary Table 5). We then reanalysed our model with the inclusion of term-born only (n = 1343) and saw only minor changes in effect estimates as compared to primary analysis (Supplementary Table 6). We then wanted to confirm our results with teacher assessments at age ten (n = 1298). The sex x BW-PGS interaction was consistent with results from our primary analysis, suggesting that a decrease in BW-PGS resulted in increased CBCL scores for males as compared to females (supplementary Table 7).

As an additional sensitivity analysis, we reanalysed our models using a BW-PGS developed based on variants identified from the BW GWAS published in 2016 (BW-PGS2) [40]. The BW-PGS explained 36 % of the variance in the BW-PGS2 and the BW-PGS2 had a modestly better predictive value for measured BW (100 g pr 1-SD increase) (see suppl. Fig 1). We saw directionally similar results compared to the BW-PGS results, with effect estimates that were modestly reduced (Supplementary Table 8). Adding PCs did not influence effect estimates (Supplementary Table 9).

Finally we investigated whether BW phenotype-genotype interactions affected behaviour. BW-PGS and BW-ENV (measured BW adjusted for BW-PGS) had significant interactions when modelling aggression problems in males (Table 2), suggesting that higher PGS in the presence of lower BW-ENV resulted in increased scores of aggression as compared to higher PGS in the presence of higher BW-ENV (Figure 4). This result was consistent when using BW-PGS2. We did not find similar interactions for attention- or social problems (p-values > 0.05) (data not shown).

**Figure 4.**
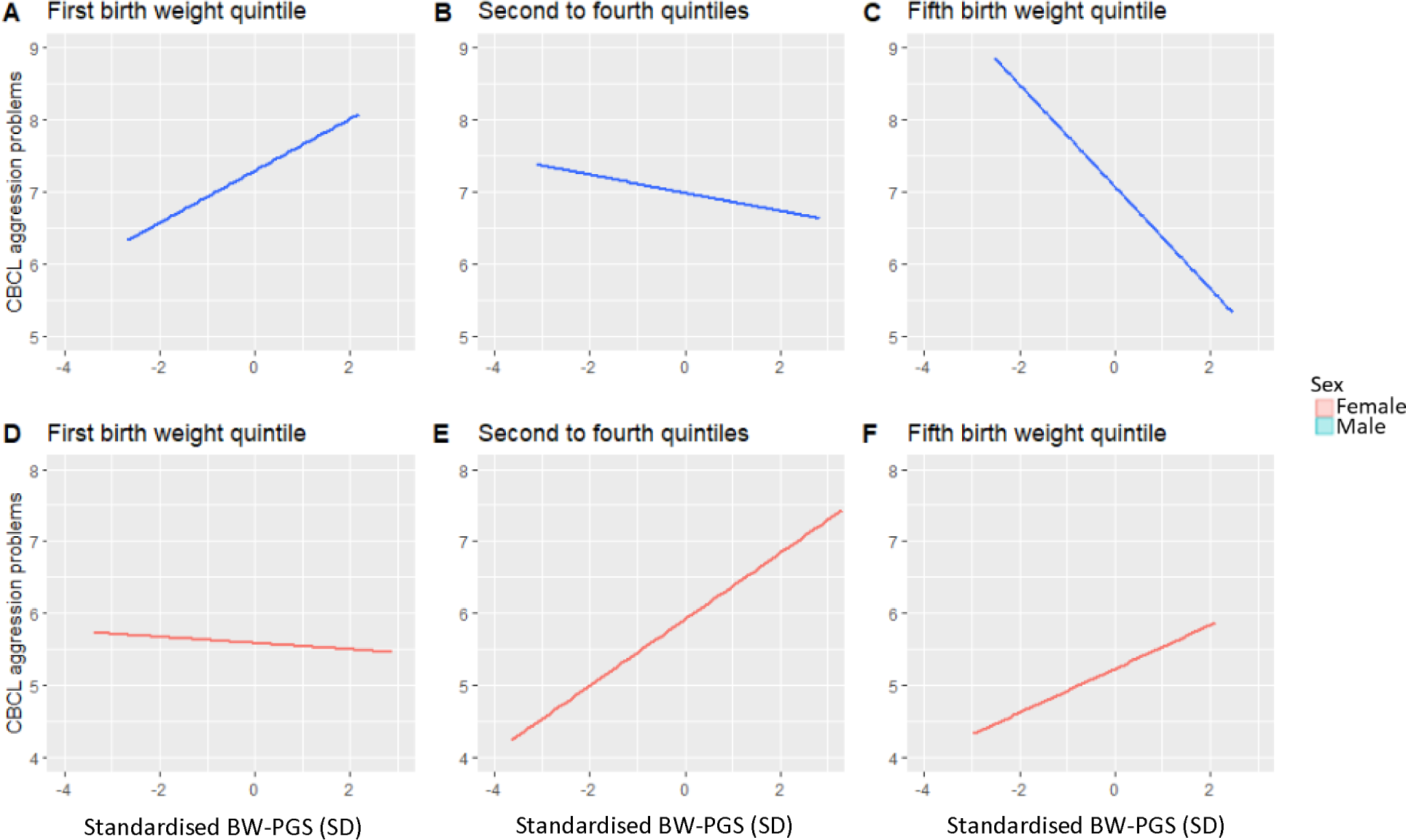
Birth weight phenotype genotype interactions for aggression. Illustration that in phenotypically smaller males (first birthweight quintile), a higher BW genotype increases CBCL aggression problem raw scores, whereas in phenotypically larger males (fifth birth weight quintile) a lower BW genotype increases aggression scores. This changing effect was statistically significant in males (**A-C**) but not females (**D-F**). Because of repeated measures no

**Table 2.**
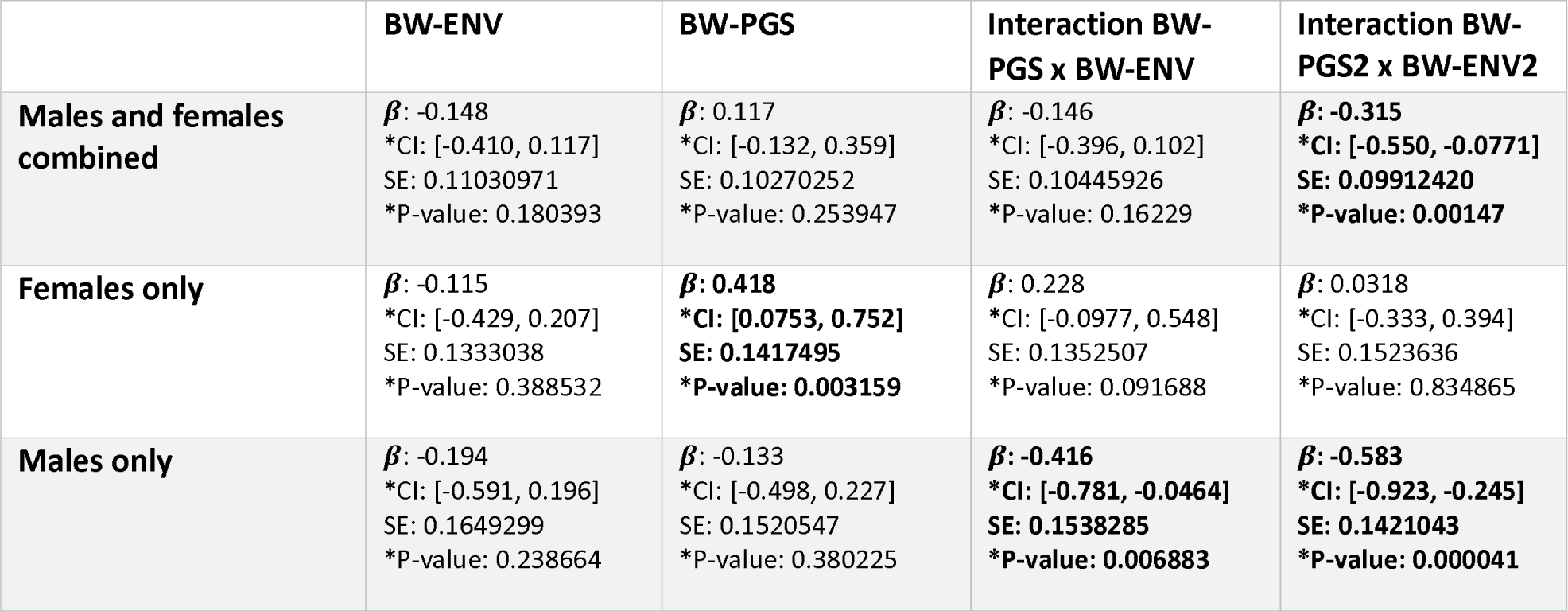
Birth weight phenotype-genotype interaction for aggression. The interaction between measured birth weight and birth weight polygenic score in linear mixed effects model. Models were made unstratified for sex and with males and females separately. **BW-PGS** = Primary birth weight polygenic score used for analysis, **BW-PGS2** = Secondary birth weight polygenic score made from an older GWAS, **BW-ENV** = measured BW adjusted for BW-PGS, **BW-ENV2** = measured BW adjusted for BW-PGS2. * p-value was calculated based on the z-statistic from the bootstrapped standard error, and the CI represents the 98.3% confidence interval.

## Discussion

Using repeated behavioural assessments across childhood and adolescence, we demonstrate two different interactions with BW: the first, a sex by genotype interaction shows increased aggression problems in males as compared to females with lower BW genotype; the second, a genotype-by-phenotype interaction, shows differential effects on aggression depending on the discrepancy between genetically determined and environmentally achieved BW, suggesting that it is this “mismatch” that drives poor outcomes.

The present study is consistent with the established epidemiology of behavioural sex differences with higher CBCL scores in males for the behaviours of interest [27]. The sex-by-phenotype interactions are consistent with results from most previous studies and suggests a male vulnerability at lower BW phenotype [13, 15].

We found no evidence for a sex-by-genotype interaction in the development of social problems drawing the causal nature of this association into question. For attention problems, a non-significant signal suggested a sex-by-genotype interaction could be present but this was inconclusive. This is surprising given the reproducible association between BW phenotype and ADHD [41] along with the sex interactions for the BW phenotype reported here and elsewhere [13, 15]. Our null-finding could reflect insufficient power to detect the difference but alternatively, it suggests that genetic determinants of BW are not important for the association between BW phenotype and ADHD, as proposed elsewhere [41].

The a priori hypothesis of a sex-by-genotype interaction causing increased CBCL scores at lower ranges of BW-PGS for males compared to females was confirmed when modelling aggression problems; however, the effects size of the sex interaction was unexpectedly large. A 1-SD increase in BW-PGS corresponded to 88.9 g change in measured BW (which had a 1-SD of approximately 580 g). If measured BW mediates the sex difference resulting from BW-PGS, this would mean that an unconfounded 1-SD decrease of measured BW would cause a 3.8-point higher score in aggression problems in males as compared to females. In the ABCD cohort, a one kg change in measured BW caused a 0.38-point change in aggression problem scores for males as compared to – more than a tenfold smaller difference compared to what we saw in our study. We saw sex interactions of similar magnitude in the teacher sensitivity analysis at age ten. Because of this large effect size, we created a second genetic instrument from an older GWAS, the BW-PGS2, which yielded results consistent with primary analysis.

When examining males and females separately another surprising finding emerged. Whereas sex differences associated with BW phenotype resulted from a detrimental effect in males at the lower range, the sex difference seen in aggression problems was driven by a protective effect in females at the lower BW range. This protective female effect was also present for attention problems. This change in what drove the interaction (from males doing worse at lower BW to females doing better at lower BW) made straightforward mediation analysis of the BW genotype through BW phenotype invalid (no exposure-outcome or mediator-outcome association, respectively) suggesting, that different causal pathways are at play. Environmental determinants of measured BW, such as smoking, have different effects on BW phenotype in males and females, so this discrepancy could simply reflect confounding [42]. Alternatively, we propose that a failure to meet your fetal growth potential has a detrimental effect in both males and females but at the population level, the consequence for females is masked by a protective effect from a lower BW genotype. This would also explain the discrepant results for genetically diverse populations vs populations with more genetically similar controls [20]. The protective effect of BW-genotype in females and the detrimental effect of BW phenotype in males is in stark contrast with Murray et al. who reported a sex-by-phenotype interaction with a female vulnerability to attention problems at lower BW [14]. Possible explanations for this discrepancy are the age four CBCL assessment, less favourable sociodemographic characteristics and a lower range of phenotypic BW in the PELOTAS cohort [43]. Alternatively, measured BW could interact with the diverging genetics of ancestrally diverse populations (Brazilian and Australian) to change associations between BW phenotype and attention problems as reported by Rahman et al [44].

We point out that our genotype by phenotype interaction was only detected post hoc, unlike our sex by genotype interaction which was postulated a priori. Again, aggression problems emerged as a trait vulnerable to a genotype-phenotype mismatch with effects isolated to the males. Our results suggest, that males with low BW phenotype are at risk of increased aggression problems only if they have a high BW genotype. This finding is supported by experimental growth restriction in primates leading to a male increase in aggression problems [11]. The reversal of effect estimates from BW-env contingent on genotype question the ability of monozygotic twin studies to uncover the full range of developmental programming as potential programming from BW discordance will depend on the shared genotype; however, this result should be seen in the light of two limitations. First, the variance of BW explained by genetics is estimated around 17 % [19] while our genetic scores explained 2-3 %; therefore, our results could represent a gene-by-gene interaction; however, the consistent effect when using the BW-PGS2 make a gene-by-gene interaction more unlikely. Second, the “BW environment” variable was generated from measured BW which is a confounded variable; however, the association was present in males only, so to bias our results the underlying confounder architecture would have to differ by both the random allocation of sex and BW-PGS. Finally, our finding of a “frail male” is consistent with animal and human experiments and has been proposed to have a placental origin as male and female placental responses to stressors are different [45–47].

The neurobiological mechanisms underlying sex-differences in aggression related to BW are not elucidated by this study or to our knowledge any study; however, fetal neurodevelopment differs by offspring sex [48–50] and is in turn affected by BW [51]. A study from the UK biobank suggested that BW-PGS and measured BW affected adult depression through various brain regions, and that these effects differed by offspring sex [52]. Similarly, a recent study found that caudate nucleus volumes in neonates had opposite associations to polygenic scores for adult depression providing evidence for sex-dimorphic relationships between fetal development and mental health [53]. For aggression, a study of 193 children and adolescents suggested that the orbitofrontal and right anterior cingulate cortex volume had gender specific associations to aggressive behaviour. The morphology of the anterior cingulate and orbitofrontal cortex associated with low BW [51] and extreme low BW[54]. In light of our results, behavioural sex differences stemming from these effects warrant investigation.

This study has several strengths. Regressions using BW-phenotype agreed with previous authors, suggesting that our exposure-outcome associations are generalisable. As accepted with Mendelian randomisation designs, the BW-PGS should not be subject to confounding. The three behavioural outcomes were based on previous studies and tested at a conservative significance threshold. Prospective repeated assessments with a single validated instrument should increase accuracy and eliminate recall bias, and the teacher assessments provide evidence that our results are not caused by bias in parental assessments. The use of a second PGS to confirm our results, supports the validity of our inference and we saw robust results throughout the sensitivity analyses. Finally, we had a 99.3 % follow-up of the genotyped participants.

Our limitations are the primarily Caucasian ethnicity in our cohort and the age 17 cut-off for our behavioural assessment; additionally, we saw normal distributions of the genetic scores, but the cohort without genetic information had different maternal baseline characteristics, which could reduce generalisability.

In conclusion, we have used robust causal inference methods to confirm that birth weight affects behaviour in males and females differently across childhood and adolescence and our results encourage incorporation of genetic scores in future twin studies. Our results also suggest that the discrepancy between genetically-determined and environmentally-achieved BW, i.e. “mismatch”, could be the driver of pathology rather than absolute BW per se but this will need to be confirmed in further studies. Finally, basic research into mechanisms underlying sex differences and gene-environment interactions in behavioural outcomes from environmental and genetic determinants of BW are needed to facilitate possible interventions.

## Data Availability

The data sets generated during and/or analyzed during the present study are not available. The Raine Study is committed to a high level of confidentiality of the data in line with the informed consent provided by participants. Requests for data should be directed to the Raine Study Executive.

## Acknowledgements

We would like to acknowledge the Raine Study participants and their families for their ongoing participation in the study and the Raine Study team for study co-ordination and data collection. We also thank the NHMRC for their long-term contribution to funding the study over the last 30 years. The core management of the Raine Study is funded by The University of Western Australia, Curtin University, Telethon Kids Institute, Women and Infants Research Foundation, Edith Cowan University, Murdoch University, The University of Notre Dame Australia and the Raine Medical Research Foundation. The Raine Study Gen2-14 year follow-up was sponsored by the NHMRC Program Grant 211912 and 003209. The Raine Study Gen2-17 year follow-up was sponsored by the NHMRC Program Grant 353514. DNA or GWAS data from the Gen2-17 year follow-up was sponsored by the NHMRC (Palmer et al, ID 572613; Beilin et al, ID 403981; Huang et al, ID 1059711) and the Canadian Institutes of Health Research - CIHR (Lye et al, MOP-82893). The Pawsey Supercomputing Centre provided computation resources to carry out analyses required with funding from the Australian Government and the Government of Western Australia.

