## Supplementary for "Sex-specific effects of Birth Weight on Longitudinal Behavioural Outcomes; a Mendelian Randomisation Approach using Polygenic Scores"

Supplementary materials

### Supplementary methods

#### Genetics data

The Raine Study Gen2 participants were genotyped at the Centre for Applied Genomics, Toronto, Canada. Quality control (QC) of the Genome-Wide-Association-Study (GWAS) genotyped data were performed as per standard protocol. In brief, a total of 1593 Raine Study Gen2 participants were genotyped on an Illumina 660 Quad Array, which included 657,366 genetic variants, consisting of ~ 560,000 single-nucleotide-polymorphisms (SNPs) and ~ 95,000 copy number variants (CNVs). Plate controls and replicates with a higher proportion of missing data were excluded before individuals were assessed for low genotyping success (> 3% missing), excessive heterozygosity, gender discrepancies between the core data and genotyped data, and cryptic relatedness (π > 0.1875, in between second- and third-degree relatives—e.g. between half-siblings and cousins). At the SNP level, the SNP data were cleaned using plink[1] following the Wellcome Trust Case—Control Consortium protocol[2]. The exclusion criteria for SNPs included: Hardy–Weinberg-Equilibrium p < 1E-06; call-rate < 95%; minor-allele frequency < 1%; and SNPs of possible strand ambiguity (i.e. A/T and C/G SNPs). The cleaned GWAS data were phased and imputed using shapeit[3] and Minimac3 [4] on the Michigan Imputation Server [4] across the 22-autosomes and X-chromosome against the Haplotype Reference Consortium reference panel [5]. A total of 1494 individuals with 518,288 SNPs remained after genotype QC; imputation resulted in 39,131,578 and 1,273,927 SNPs across the 22-autosomes and X-chromosome, respectively. Principal components (PCs) analysis was carried out, using SMARTPCA from v.3.0 of EIGENSOFT[6] , on the cleaned genotyped data, where PCs were generated for purposes of adjusting for population stratification in all genetic analyses.

### Supplementary Figure 1. Effect of the second polygenic score BW-PGS2

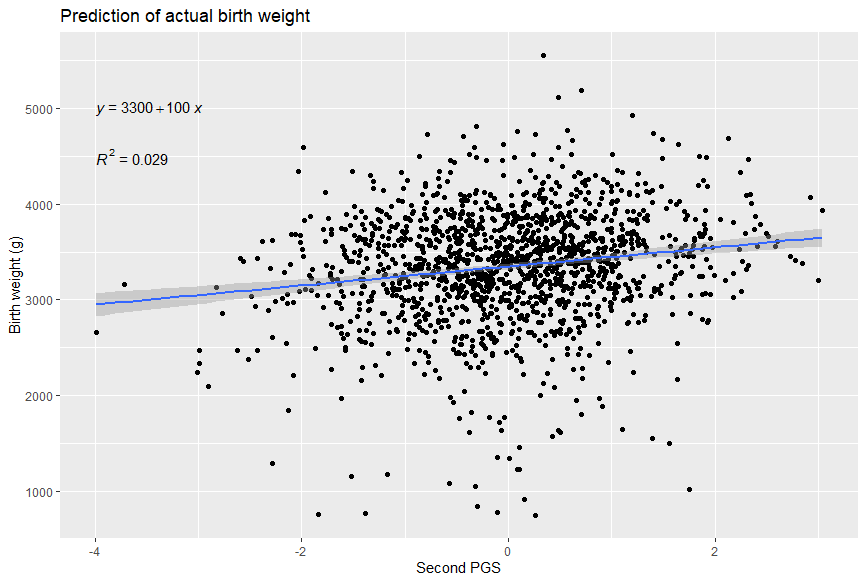

**a**

BW-PGS2 (SD)

BW-PGS (SD)

**b**

Birth weight (g)

BW-PGS2 (SD)

Supplementary Figure 1: The correlation between the BW-PGS used in primary analysis, and the second BW-PGS (BW-PGS2) used in sensitivity analysis (**a**) and the predictive value of the BW-PGS (**b**) for measured birth weight in our cohort in males and females combined. **BW-PGS**: Birth weight polygenic score.

### Supplementary Table 1. Behaviour- and birth weight variables

| Baseline information in Raine Study participants with genetic information and 1 or more Child behaviour checklist scores from ages 5 – 17 (n = 1484) | | | |
| --- | --- | --- | --- |
|  | **Female** (n = 722) | **Male** (n = 762) | **P-value*** |
| **Measured birth weight (g)** | | | < 0.0001 |
| Mean (SD) | 3291.9 (±574.7) | 3404.2 (±588.7) |  |
| Missing | 0 (0%) | 1 (0.1%) |  |
| **Polygenic score for birth weight (normalised)** | | | 0.84 |
|  | 0.0 (±1.0) | 0.0 (±1.0) |  |
| **CBCL Aggression problems age 5** | | | < 0.0001 |
| Mean (SD) | 7.6 (±5.6) | 9.3 (±6.6) |  |
| Missing | 59 (8.2%) | 62 (8.1%) |  |
| **CBCL Aggression problems age 8** | | | < 0.0001 |
| Mean (SD) | 6.4 (±5.7) | 8.3 (±7.0) |  |
| Missing | 65 (9.0%) | 72 (9.4%) |  |
| **CBCL Aggression problems age 10** | | | < 0.0001 |
| Mean (SD) | 5.2 (±5.1) | 7.2 (±6.5) |  |
| Missing | 65 (9.0%) | 53 (7.0%) |  |
| **CBCL Aggression problems age 14** | | | 0.043 |
| Mean (SD) | 5.3 (±5.8) | 5.8 (±6.1) |  |
| Missing | 74 (10.2%) | 79 (10.4%) |  |
| **CBCL Aggression problems age 17** | | | 0.6 |
| Mean (SD) | 3.7 (±4.7) | 3.9 (±5.0) |  |
| Missing | 220 (30.5%) | 225 (29.5%) |  |
| **CBCL Attention problems age 5** | | | < 0.0001 |
| Mean (SD) | 2.5 (±2.6) | 3.4 (±2.9) |  |
| Missing | 59 (8.2%) | 62 (8.1%) |  |
| **CBCL Attention problems age 8** | | | < 0.0001 |
| Mean (SD) | 2.4 (±2.8) | 3.6 (±3.4) |  |
| Missing | 65 (9.0%) | 72 (9.4%) |  |
| **CBCL Attention problems age 10** | | | < 0.0001 |
| Mean (SD) | 1.9 (±2.7) | 3.2 (±3.4) |  |
| Missing | 65 (9.0%) | 53 (7.0%) |  |
| **CBCL Attention problems age 14** | | | < 0.0001 |
| Mean (SD) | 1.9 (±2.5) | 2.7 (±3.1) |  |
| Missing | 74 (10.2%) | 79 (10.4%) |  |
| **CBCL Attention problems age 17** | | | 0.001 |
| Mean (SD) | 1.5 (±2.2) | 2.0 (±2.7) |  |
| Missing | 220 (30.5%) | 225 (29.5%) |  |
| **CBCL Social problems age 5** | | | 0.026 |
| Mean (SD) | 1.5 (±1.7) | 1.7 (±1.8) |  |
| Missing | 59 (8.2%) | 62 (8.1%) |  |
| **CBCL Social problems age 8** | | | 0.3 |
| Mean (SD) | 1.5 (±1.9) | 1.7 (±2.1) |  |
| Missing | 65 (9.0%) | 72 (9.4%) |  |
| **CBCL Social problems age 10** | | | 0.001 |
| Mean (SD) | 1.4 (±2.0) | 1.8 (±2.1) |  |
| Missing | 65 (9.0%) | 53 (7.0%) |  |
| **CBCL Social problems age 14** | | | 0.12 |
| Mean (SD) | 1.1 (±1.8) | 1.3 (±1.9) |  |
| Missing | 74 (10.2%) | 79 (10.4%) |  |
| **CBCL Social problems age 17** | | | 0.6 |
| Mean (SD) | 0.7 (±1.4) | 0.7 (±1.4) |  |
| Missing | 220 (30.5%) | 225 (29.5%) |  |
| * P-value calculated using Wilcoxon-signed rank test | | | |

### Supplementary Table 2. Baseline demographics males vs females

| Demographics for Raine Study participants with genetic data and at least one CBCL score from age 5 - 17 (n = 1484) | | |
| --- | --- | --- |
|  | **Female** (n = 722) | **Male** (n = 762) |
| **Maternal age at birth (years)** | | |
| Mean (SD) | 28.8 (±5.9) | 28.9 (±5.7) |
| Missing | 11 (1.5%) | 4 (0.5%) |
| **Income level†** | | |
| Below 12.000 AUD | 89 (12.3%) | 90 (11.8%) |
| Above 12.000 AUD | 589 (81.6%) | 641 (84.1%) |
| Missing | 44 (6.1%) | 31 (4.1%) |
| **Maternal body mass index (kg/m^2)** | | |
| Mean (SD) | 22.5 (±4.3) | 22.5 (±4.3) |
| Missing | 11 (1.5%) | 4 (0.5%) |
| **Maternal race** | | |
| European descent | 687 (95.2%) | 737 (96.7%) |
| Other | 24 (3.3%) | 21 (2.8%) |
| Missing | 11 (1.5%) | 4 (0.5%) |
| **Maternal level of education** | | |
| < 12 years | 350 (48.5%) | 351 (46.1%) |
| > 12 years | 361 (50.0%) | 407 (53.4%) |
| Missing | 11 (1.5%) | 4 (0.5%) |
| **Diabetes or hypertension in pregnancy** | | |
| absent | 585 (81.0%) | 620 (81.4%) |
| present | 126 (17.5%) | 138 (18.1%) |
| Missing | 11 (1.5%) | 4 (0.5%) |
| **Gestational age at birth (weeks)** | | |
| Mean (SD) | 38.8 (±2.2) | 38.8 (±2.1) |
| Missing | 1 (0.1%) | 1 (0.1%) |
| **Smoking in pregnancy** | | |
| Non-smoker | 510 (70.6%) | 576 (75.6%) |
| Smoker | 153 (21.2%) | 140 (18.4%) |
| Missing | 59 (8.2%) | 46 (6.0%) |
| **Any maternal psychiatric illness** | | |
| Absent | 697 (96.5%) | 738 (96.9%) |
| present | 14 (1.9%) | 20 (2.6%) |
| Missing | 11 (1.5%) | 4 (0.5%) |
| **Maternal alcohol consumption in first three months** | | |
| Any alcohol | 342 (47.4%) | 388 (50.9%) |
| Never | 369 (51.1%) | 370 (48.6%) |
| Missing | 11 (1.5%) | 4 (0.5%) |
| †Family income was recorded in 1990, explaining the low absolute monetary level. | | |

### Supplementary Table 3. Analytic vs excluded cohort

| Demographics for analytic cohort vs excluded Raine Study participants (total n = 2868) | | |
| --- | --- | --- |
|  | **Excluded cohort** (n = 1384) | **Analytic cohort** (n = 1484) |
| **Maternal age at birth (years)** | | |
| Mean (SD) | 27.2 (±6.0) | 28.9 (±5.8) |
| Missing | 57 (4.1%) | 15 (1.0%) |
| **Female fetal sex** |  |  |
|  | 692 (50.0%) | 722 (48.7%) |
| **Income level†** | | |
| Below 12.000 AUD | 292 (21.1%) | 179 (12.1%) |
| Above 12.000 AUD | 937 (67.7%) | 1,230 (82.9%) |
| Missing | 155 (11.2%) | 75 (5.1%) |
| **Maternal body mass index (kg/m^2)** | | |
| Mean (SD) | 22.2 (±4.4) | 22.5 (±4.3) |
| Missing | 50 (3.6%) | 15 (1.0%) |
| **Maternal race** | | |
| European descent | 1,048 (75.7%) | 1,424 (96.0%) |
| Other | 287 (20.7%) | 45 (3.0%) |
| Missing | 49 (3.5%) | 15 (1.0%) |
| **Maternal level of education** | | |
| < 12 years | 747 (54.0%) | 701 (47.2%) |
| > 12 years | 588 (42.5%) | 768 (51.8%) |
| Missing | 49 (3.5%) | 15 (1.0%) |
| **Diabetes or hypertension in pregnancy** | | |
| absent | 1,126 (81.4%) | 1,205 (81.2%) |
| present | 209 (15.1%) | 264 (17.8%) |
| Missing | 49 (3.5%) | 15 (1.0%) |
| **Gestational age at birth (weeks)** | | |
| Mean (SD) | 38.5 (±2.6) | 38.8 (±2.2) |
| Missing | 9 (0.7%) | 2 (0.1%) |
| **Smoking in pregnancy** | | |
| Non-smoker | 819 (59.2%) | 1,086 (73.2%) |
| Smoker | 351 (25.4%) | 293 (19.7%) |
| Missing | 214 (15.5%) | 105 (7.1%) |
| **Any maternal psychiatric illness** | | |
| Absent | 1,303 (94.1%) | 1,435 (96.7%) |
| present | 32 (2.3%) | 34 (2.3%) |
| Missing | 49 (3.5%) | 15 (1.0%) |
| **Maternal alcohol consumption in first three months** | | |
| Any alcohol | 549 (39.7%) | 730 (49.2%) |
| Never | 786 (56.8%) | 739 (49.8%) |
| Missing | 49 (3.5%) | 15 (1.0%) |

†Family income was recorded in 1990, explaining the low absolute monetary level.

### Supplementary Table 4. Measured birthweight

|  | Aggression problems | | Attention problems | | Social problems | |
| --- | --- | --- | --- | --- | --- | --- |
|  | **Model 1** | **Model 2** | **Model 1** | **Model 2** | **Model 1** | **Model 2** |
| **Main effect (normalized birth weight (SD))** | 𝜷: -0.0618  *CI: [-0.313, 0.191]  SE: 0.105  *P-value: 0.558 | NA | 𝜷: -0.0954  *CI: [-0.216, 0.0256]  SE: 0.0505  *P-value: 0.059 | NA | 𝜷: -0.0604  *CI:[-0.135, 0.014]  SE: 0.0312  *P-value: 0.0532 | NA |
| **Female effect** | NA | 𝜷: -0.0460941  *CI: [-0.370, 0.286]  SE: 0.1372888  *P-value: 0.737 | NA | 𝜷: -0.0273968  *CI: [-0.174, 0.111]  SE: 0.05864  *P-value: 0.64 | NA | 𝜷: -0.0046495  *CI: [-0.113, 0.101]  SE: 0.04478380  *P-value: 0.917 |
| **Male effect** | NA | 𝜷: -0.198  *CI: [-0.601, 0.199]  SE: 0.168  *P-value: 0.238 | NA | 𝜷: -0.250  *CI:[-0.43, -0.0696]  SE: 0.0757  *P-value: 0.00096 | NA | 𝜷: -0.128  *CI:[-0.232, -0.024]  SE: 0.0436  *P-value: 0.0033 |
| **Interaction on female effect from male sex** | NA | 𝜷: -0.1519129  *CI: [-0.673, 0.345]  SE: 0.2133396  *P-value: 0.476 | NA | 𝜷: -0.2224331  *CI: [-0.454, 0.003]  SE: 0.0955  *P-value: 0.0199 | NA | 𝜷: -0.1235780  *CI: [-0.271, 0.027]  SE: 0.06245463  *P-value: 0.0479 |

Supplementary Table 4: Regressing the measured birthweight on scores of aggression, attention and social problems in males and females together and with the inclusion of an age interaction the regression output for our linear mixed effects model.

* p-value was calculated based on the z-statistic from the bootstrapped standard error, and the CI represents the 98.3% confidence interval.

### Supplementary Table 5. Aggression with age two assessment

|  | Model 1 | Model 2 |
| --- | --- | --- |
| **Main effect** | 𝜷: 0.141  *CI: [-0.119, 0.391]  SE: 0.104  *P-value: 0.187476 | NA |
| **Female effect** | NA | 𝜷: 0.511  *CI: [0.164, 0.849 ]  SE: 0.14342248  *P-value: 0.00037 |
| **Male interaction** | NA | 𝜷: -0.734  *CI: [-1.25, -0.237 ]  SE: 0.213  *P-value: 0.000561 |

### Supplementary Table 6. Term born only

Supplementary Table 5: Sensitivity analysis with inclusion of the age two preschool assessment showing the regression output for our linear mixed effects model.

* p-value was calculated based on the z-statistic from the bootstrapped standard error, and the CI represents the 98.3% confidence interval.

|  | Aggression Problems | Attention Problems | Social Problems |
| --- | --- | --- | --- |
| **Female effect** | 𝜷: 0.387  *CI: [0.011, 0.762 ]  SE: 0.157  *P-value: 0.014 | 𝜷: 0.142  *CI: [-0.0318, 0.311]  SE: 0.072  *P-value: 0.048 | 𝜷: 0.0521  *CI: [-0.085, 0.182]  SE: 0.056  *P-value: 0.35151 |
| **Male interaction** | 𝜷: -0.646  *CI: [-1.19, -0.114 ]  SE: 0.226  *P-value: 0.0042 | 𝜷: -0.148  *CI: [-0.389, 0.0982]  SE: 0.102  *P-value: 0.148 | 𝜷: -0.0158  *CI: [-0.186, 0.152 ]  SE: 0.071  *P-value: 0.823 |

Supplementary Table 6: Sensitivity analysis with term-born only showing the regression output for our linear mixed effects model.

* p-value was calculated based on the z-statistic from the bootstrapped standard error, and the CI represents the 98.3% confidence interval.

### Supplementary Table 7. Teacher assessments

|  | Aggression Problems | Attention Problems | Social Problems |
| --- | --- | --- | --- |
| **Female effect** | 𝜷: 0.113  *CI: [-0.349, 0.573]  SE: 0.193  *P-value: 0.559 | 𝜷: 0.138  *CI: [-0.432, 0.7]  SE: 0.237  *P-value: 0.561 | 𝜷: -0.029  *CI: [-0.32, 0.263]  SE: 0.122  *P-value: 0.813 |
| **Male interaction** | 𝜷: -0.792  *CI: [-1.625, 0.079]  SE: 0.357  *P-value: 0.026 | 𝜷: -0.785  *CI: [-1.705, 0.177]  SE: 0.394  *P-value: 0.0463 | 𝜷: -0.15  *CI: [-0.538, 0.239]  SE: 0.163  *P-value: 0.357 |

### Supplementary Table 8. Sex interactions when using BW-PGS2

Supplementary Table 7: supplementary analysis using teacher assessments at age ten showing the regression output for our linear model after bootstrapping standard errors and confidence intervals.

* p-value was calculated based on the z-statistic from the bootstrapped standard error, and the CI represents the 98.3% confidence interval.

|  | Attention Problems | Aggression Problems | Social Problems |
| --- | --- | --- | --- |
| **Female effect** | 𝜷: 0.125  *CI: [-0.0394, 0.288]  SE: 0.069  *P-value: 0.068 | 𝜷: 0.292  *CI: [-0.0618, 0.647]  SE: 0.148  *P-value: 0.049 | 𝜷: 0.0552  *CI: [-0.0574, 0.167]  SE: 0.047  *P-value: 0.24 |
| **Male interaction** | 𝜷: -0.139  *CI: [-0.370, 0.101]  SE: 0.099  *P-value: 0.160 | 𝜷: -0.510  *CI: [-1.01, -0.00477]  SE: 0.210  *P-value: 0.0149 | 𝜷: -0.0496  *CI: [-0.195, 0.100]  SE: 0.062  *P-value: 0.422 |

Supplementary Table 8: Sensitivity analysis using an older polygenic score with regression output for our linear mixed effects model. **BW-PGS2** = Secondary BW polygenic score made from an older GWAS.

* p-value was calculated based on the z-statistic from the bootstrapped standard error, and the CI represents the 98.3% confidence interval.

### Supplementary Table 9. Including Principal components

|  | Attention Problems | Aggression Problems | Social Problems |
| --- | --- | --- | --- |
| **Female effect** | 𝜷: 0.181  *CI: [0.0213, 0.335]  SE: 0.0657  *P-value: 0.006 | 𝜷: 0.415  *CI: [0.0711, 0.747]  SE: 0.1417  *P-value: 0.003 | 𝜷: 0.0756  *CI: [-0.0458, 0.194]  SE: 0.0502  *P-value: 0.1318 |
| **Male interaction** | 𝜷: -0.190  *CI: [-0.421, 0.0411]  SE: 0.0968  *P-value: 0.049 | 𝜷: -0.5887  *CI: [-1.08, -0.0804]  SE: 0.2096  *P-value: 0.005 | 𝜷: -0.0420  *CI: [-0.202, 0.116]  SE: 0.0666  *P-value: 0.528 |

Supplementary Table 9: Sensitivity analysis including principal components to investigate potential confounding from population stratification. Adjusted for sex, age, and principal components with clustering at the participant level

* p-value was calculated based on the z-statistic from the bootstrapped standard error, and the CI represents the 98.3% confidence interval.
